# The association between HIV treatment interruptions and viral suppression during the early treatment period: retrospective cohort study in South Africa

**DOI:** 10.64898/2026.08.20.26360935

**Authors:** Mariet Benade, Mhairi Maskew, Nyasha Mutanda, Nancy Scott, Allison Morgan, Vinolia Ntjikelane, Linda Sande, Lufuno Malala, Musa Manganye, Brooke Nichols, Sydney Rosen

## Abstract

**Background:** The first six months after antiretroviral therapy (ART) initiation for HIV is a high-risk period for treatment interruptions that may compromise viral suppression (VS). Recent research in South Africa suggests that >40% of patients interrupt care for >28 days during the early treatment period. The quantitative association between early treatment interruptions and VS at 6 and 12 months remains unclear.

**Methods:** We enrolled adults (≥18 years) initiating ART from 1 January 2018 to 7 November 2024 with at least 14 months’ followup in South Africa’s national ART database (TIER.Net) from 24 public sector facilities in four provinces. Engagement in care during months 0-6 and 7-12 was classified as continuous (no interruptions >28 days), cyclical (at least one interruption >28 days but returned to care within follow up period), or disengaged (>28 days late without return), based on completed and scheduled visit dates. Modified Poisson regression was used to estimate adjusted risk ratios (aRRs) for VS (<50 copies/mL), adjusting for age, sex, initiation year, regimen, engagement pattern, and baseline CD4 count.

**Findings:** Among 57,553 participants (66% female; median age 33 years), 49% and 42% were continuously engaged at 6 and 12 months, respectively; 22% and 17% were cyclically engaged at the same time points. 54% of continuously engaged participants achieved 6-month VS compared with 34% of those with cyclical engagement (aRR 1.60 95% CI 1.55-1.64). At 12 months, 56% of continuously engaged individuals and 40% of those cyclically engaged were suppressed (aRR 1.38 95% CI 1.34-1.42). VS was also associated with dolutegravir-based regimens, later ART initiation year, baseline CD4 count ≥200 cells/uL, female sex, and older age.

**Interpretation:** Even relatively brief treatment interruptions during the first year of ART were associated with substantially lower viral suppression. Preventing early interruptions should remain a programmatic priority to improve treatment outcomes.

**Funding:** Funding for this study was provided by the Gates Foundation under INV-031690 to Boston University.

## INTRODUCTION

Sustained viral load suppression remains the ultimate goal of national HIV treatment programs, with success measured as the proportion of all people living with HIV who have suppressed viral loads (1). Viral suppression benefits both individual health, by reducing the risk of HIV-related morbidity and mortality, and population health, by diminishing the likelihood of HIV transmission(2). Sustaining viral suppression requires, in turn, maintaining uninterrupted engagement in HIV care and a high level of medication adherence among those who have initiated antiretroviral therapy (ART). Continuous engagement during the early treatment period (initial six months after initiation) is particularly important, as this is the interval when clients establish their care journey and are first expected to achieve viral suppression. Continuity of care during this period is associated with a reduced time to viral suppression and a lower cumulative viral burden(3). Conversely, missed clinic visits during the early treatment period have been linked to delays in achieving viral suppression(4–6).

Despite the value to patients of maintaining continuity of care, treatment interruptions are common. Multiple studies have documented that the risk of interruption and/or disengagement from care is highest during the early period, when clients are not yet established in care. Recent studies report substantial losses during this time, with disengagement rates at 6 months in sub-Saharan Africa averaging 26% (7). A recent study in South Africa found that only 57% of clients remained continuously engaged in care (no interruptions >28 days) at 6 months after treatment initiation, with most of the 16% who disengaged fully during this period doing so immediately after the initiation visit (5). Other data suggest that nearly half of clients—if not more—presenting for “initiation” of ART are, in fact, returning to care after a treatment interruption(8,9). These figures suggest that the “cyclical engager”(10)–a client who experiences one or more periods of treatment interruption but does not fully disengage from care– represents an increasing share of the population on ART. Understanding the relationship between viral suppression and patterns of engagement, interruption, and disengagement from HIV care during the early treatment period is thus essential for optimizing both individual and programmatic treatment outcomes (5).

If cyclical enagagement is indeed the new norm in HIV care programs, and this pattern of engagement adversely affects viral suppression, then service delivery models should be amended to account for this phenomenon. Limited efforts to date, however, have directly quantified the associations between different patterns of engagement in care and HIV viral suppression during the early treatment period. Building on previous work describing patterns of engagement among clients in the first year after ART initiation in South Africa(5), we quantify the association between pattern of engagement in HIV care and subsequent viral suppression at 6 and 12 months and explore clinical and programmatic factors associated with suppression outcomes.

## METHODS

### Study population and data

For this analysis, we used routinely collected medical record data from South Africa’s national electronic HIV/TB patient management and monitoring system, TIER.Net(11). This data source captures demographic characteristics, visit history, and laboratory test dates and results of clients accessing HIV care and treatment in public sector facilities in South Africa.

We accessed data from 24 public sector primary health care clinics, with six facilities in each of four provinces in South Africa (Mpumalanga (MP), KwaZulu-Natal (KZN), Gauteng (GP), and Eastern Cape (EC)). These facilities comprise the SENTINEL observational research network, described elsewhere(12). Analytic datasets included all observations in client TIER.Net records between 1 January 2018 and the date of the last documented visit at the time of data extraction in each province (MP: 7 January 2026; KZN: 7 January 2026: GP: 7 November 2025; EC: 21 November 2025).

The study population consisted of all adults who initiated ART at one of the 24 study facilities during the observation period. We excluded participants younger than 18 years at the time of ART initiation and those who either had no visits documented or were missing dates for when visits were scheduled to happen.

### Study outcomes

The primary outcome for this analysis was documented suppression of HIV viral load at 6 and 12 months after treatment initiation, stratified by pattern of engagement in care. In keeping with current definitions in the South African National ART guidelines and the TIER.Net system(13), we defined a 6-month viral load as any viral load test completed more than 60 days after treatment initiation and prior to day 270 after initiation. We defined a 12-month viral load as any viral load test conducted more than 270 days after initiation and prior to 425 days after initiation of ART(8). If more than one viral load test was performed for a client during either period, we used the one that was closest to day 183 for 6-month viral loads and to day 365 for 12-month viral loads. We note that because TIER.Net is not networked among facilities, unofficial (“silent”) transfers of ART clients from one facility to another are reflected in the database as loss to follow up (non-retention), and viral load tests conducted at a facility other than the initiating site would not be captured in our dataset.

We created five categories of viral load results: 1) undetectable, defined as viral load <50 copies/mL; 2) low-level viremia (LLV), for a viral load between 50 and 199 copies/mL; 3) intermediate viremia, for a viral load of 200-999; 4) unsuppressed, for a viral load >1000; and 5) “no documented result” for those who had no evidence of a viral load test being conducted or for whom there was evidence of test conducted but no result recorded (Panel 1a).

The primary exposure used in our analysis was pattern of engagement in HIV care based on visits observed during the first year on ART. To create this exposure variable, we first defined five visit types to describe the timing of individual clinic visits relative to the scheduled appointment date (Panel 1b). An initiation visit (visit type 1) was defined as the observed date of antiretroviral therapy (ART) initiation.

Subsequent visits were classified as attended as planned (visit type 2) if they occurred on or before the scheduled date; attended late ≤28 days (visit type 3) if they occurred within 28 days after the scheduled date; and attended late >28 days (visit type 4) if they occurred more than 28 days after the scheduled date. A scheduled visit not attended at all within the period of observation (visit type 5) was used to define disengagement. We note that different studies have used shorter or longer intervals to define interruptions; we used 28 days to reflect the units of scripting and dispensing antiretroviral medications in South Africa(14,15).

Finally, we defined six patterns of engagement observed in the EMR during two periods: months 0-6 after ART initiation and months 7-12 after ART initiation (Panel 1c). Participants were classified as continuously engaged if no visit was ≥ 28 days late (pattern 1). Cyclical engagement (pattern 2) was defined as having at least one visit more than 28 days late followed by documented return to care within the relevant period. Clients were considered disengaged in months 0-6 (pattern 3) or in months 7-12 (pattern 4) if they missed their scheduled visit by more than 28 days with no evidence of return within our observation period. Clients who disengaged in months 7-12 were, by definition, continuously or cyclically engaged in months 0-6. Documented transfers to other facilities (pattern 5) and deaths (pattern 6) were recorded separately for both follow-up periods. For the patterns of engagement assignments, to allow for a minimum of 14 months of potential observation after treatment initiation, we excluded individuals who initiated after 7 November 2024 in MP, 7 November 2024 in MP, 7 September 2024 in GP; or 21 September 2024 in EC. We note that the study sample in this manuscript is limited to participants with the first two patterns of engagement (continuous and cyclical); we include definitions for and report outcomes for the other patterns for completeness but do not analyze these data.

**Panel 1.**
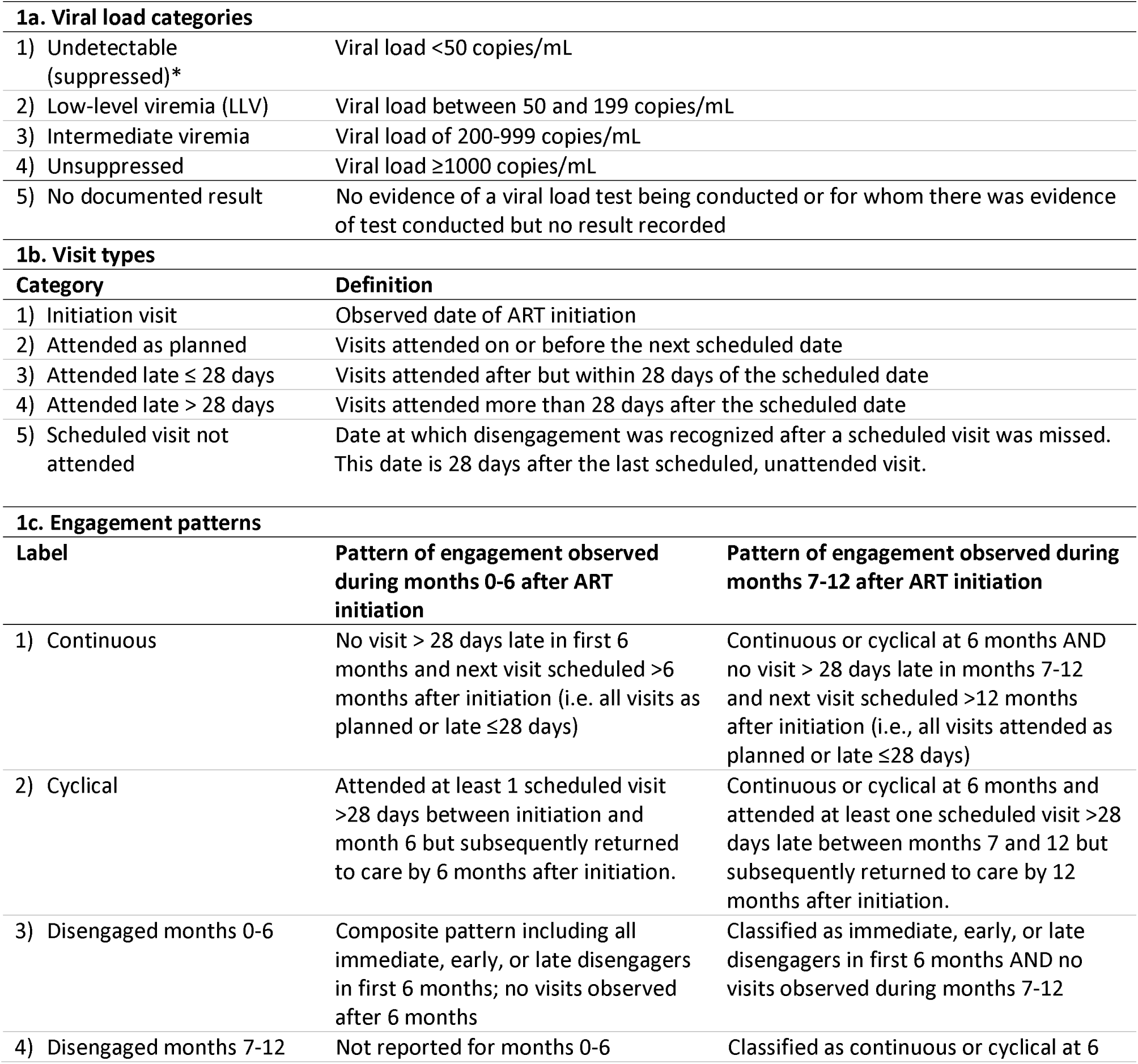
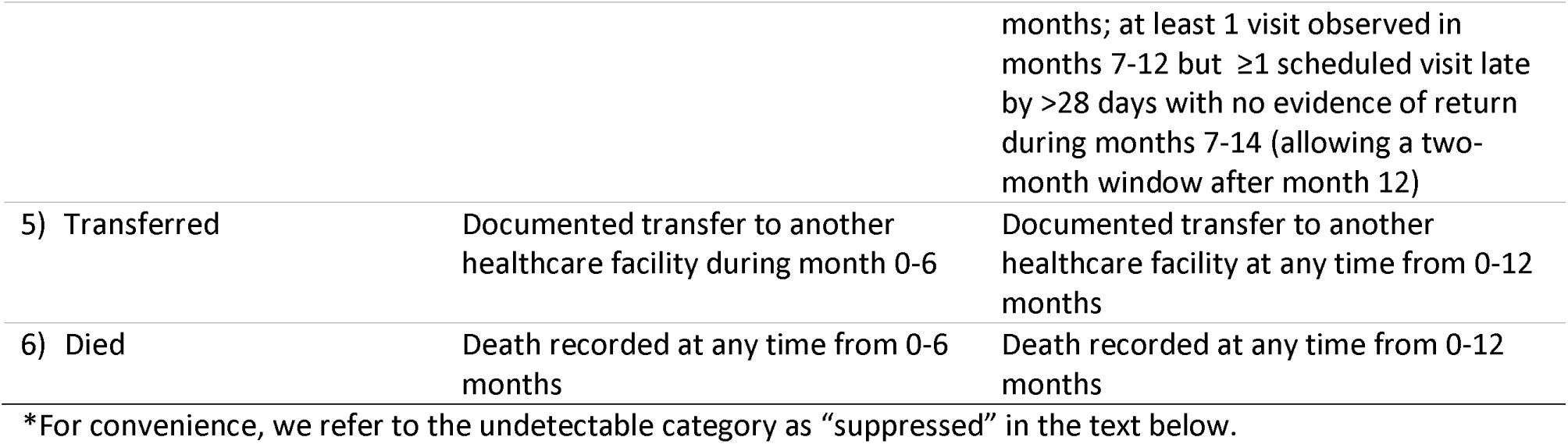
Definitions of viral loads, visit types, and engagement patterns.

### Statistical analysis

We first described characteristics of the study sample (continuously or cyclically engaged) at enrolment. Next, we estimated the patterns of engagement, timing of viral load testing, and viral suppression categories in the study sample. We then modelled the association between viral load suppression at six months (<50 copies/mL (binary outcome) and each predictor. We used a generalized linear model with a Poisson error distribution and log link function to estimate risk ratios. Robust standard errors were obtained using a heteroscedasticity-consistent sandwich variance estimator. For each predictor we first estimated the unadjusted association with viral load suppression. We then created a multivariable model including all predictors simultaneously to obtain the adjusted risk ratios and 95% confidence intervals. Predictors included year of ART initiation, drug regimen, pattern of engagement, CD4 count at baseline (for which we considered a window of 180 days prior to 7 days post ART initiation), sex, and age. Standard errors were not clustered at the facility level because the number of facilities were insufficient for reliable cluster-robust variance estimation. All analyses were conducted in R (version 4.5.2) using the sandwich (v3.1.1), lmtest (v0.9.40) and broom (v1.0.11) packages.

The drug regimens considered in this analysis were the newer regimen of tenofovir disoproxil fumarate, lamivudine, and dolutegravir (TDF/3TC/DTG) in a fixed dose combination, also known as TLD, and the previous regimen of tenofovir disoproxil fumarate, emtricitabine, and efavirenz (TDF/FTC/EFV). Due to the likelihood of collinearity between year of ART initiation and drug regimen at initiation, we also created a model that was limited to PLHIV who were initiated on a dolutegravir-containing regimen in 2020 (when about 50% of the sample were initiated on dolutegravir-based regimen) or later. This model did not include drug regimen as a predictor.

### Ethics review

The use of de-identified data for this analysis was approved by the Boston University Institutional Review Board (H-38115) and by the University of the Witwatersrand Human Research Ethics Committee (M190445). The protocol was also approved by Provincial Health Research Committees through the National Health Research Database for each study district. We received a waiver of informed consent for all study data.

## RESULTS

### Cohort characteristics

We analysed baseline initiation data for 70,930 clients who initiated ART between 1 January 2018 and 7 January 2026. After excluding people not eligible due to age (n=2,865), less than 14 months’ observation time (n=4,696), no corresponding visit data (n=44), or incomplete visit data (n=5,772), our sample included 57,553 clients who initiated HIV treatment between 1 January 2018 and 7 November 2024 (Table 1). A participant flow diagram describing cohort creation and overall 6- and 12-month outcomes is provided in Supplementary Figure 1.

**Table 1.**
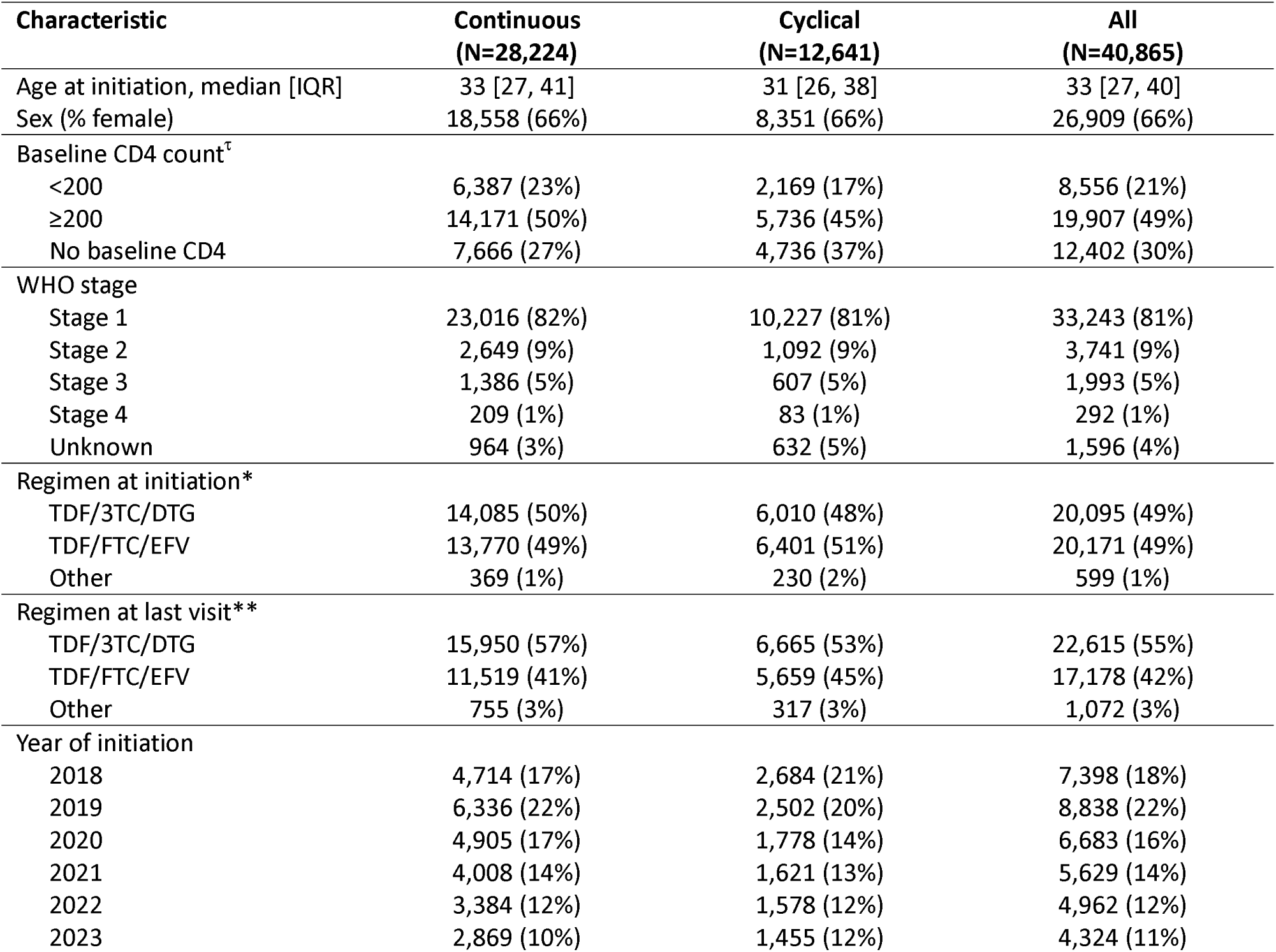
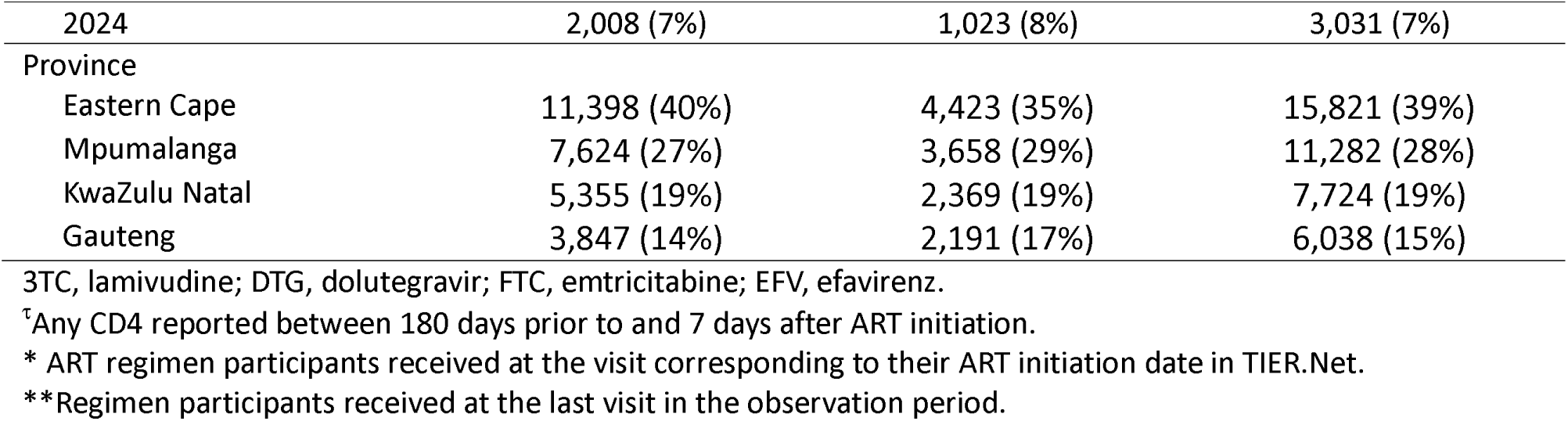
Baseline characteristics of sample population, by pattern of engagement observed during the first 6 months on ART.

Among the final enrolled sample of 57,553, 49% started on TLD (TDF/3TC/DTG) and 50% started on TDF/FTC/EFV. At the time of ART initiation, 20% had a CD4 count of less than 200 cells/mm^3^. Fewer than 6% were categorized as WHO stage 3 or stage 4, while 82% were categorized as stage 1. Among this sample, 28,224 were continuously engaged and 12,641 were cyclically engaged at 6 months, while 9,175 clients disengaged from care, 6,761 were transferred to other facilties, and 752 clients died between months 0-6. For this analysis, we restricted the study sample to the 40,865 individuals who were still engaged either continuously or cyclically at the end of the 6-month period or the 34,418 who were continuously or cyclically engaged at the end of the 12-month period. This restriction allowed us to compare continuous and cyclical engagement directly, with the presence or absence of ≥1 interruption comprising the sole difference between the cohorts.

### Changes in viral suppression over time

Figure 1 illustrates changes in viral load suppression between the first and second 6-month periods after treatment initiation, stratified by pattern of engagement among those remaining in care at 12 months (n=34,418). Among the 71% of participants who were continuously engaged (n=24,334), 54% were undetectable at 6 months, 18% had low level viremia, 4% had intermediate viremia, 5% were unsuppressed, and 19% had no viral load result. At 12 months, suppression increased to 56%, low level viremia declined to 13%, intermediate viremia was 3%, and unsuppression remained at 4%, while 25% had no 12-month viral load result. Among those suppressed at 6 months, 65% remained suppressed at 12 months, while among those with LLV at 6 months, 48% achieved suppression by 12 months.

**Figure 1.**
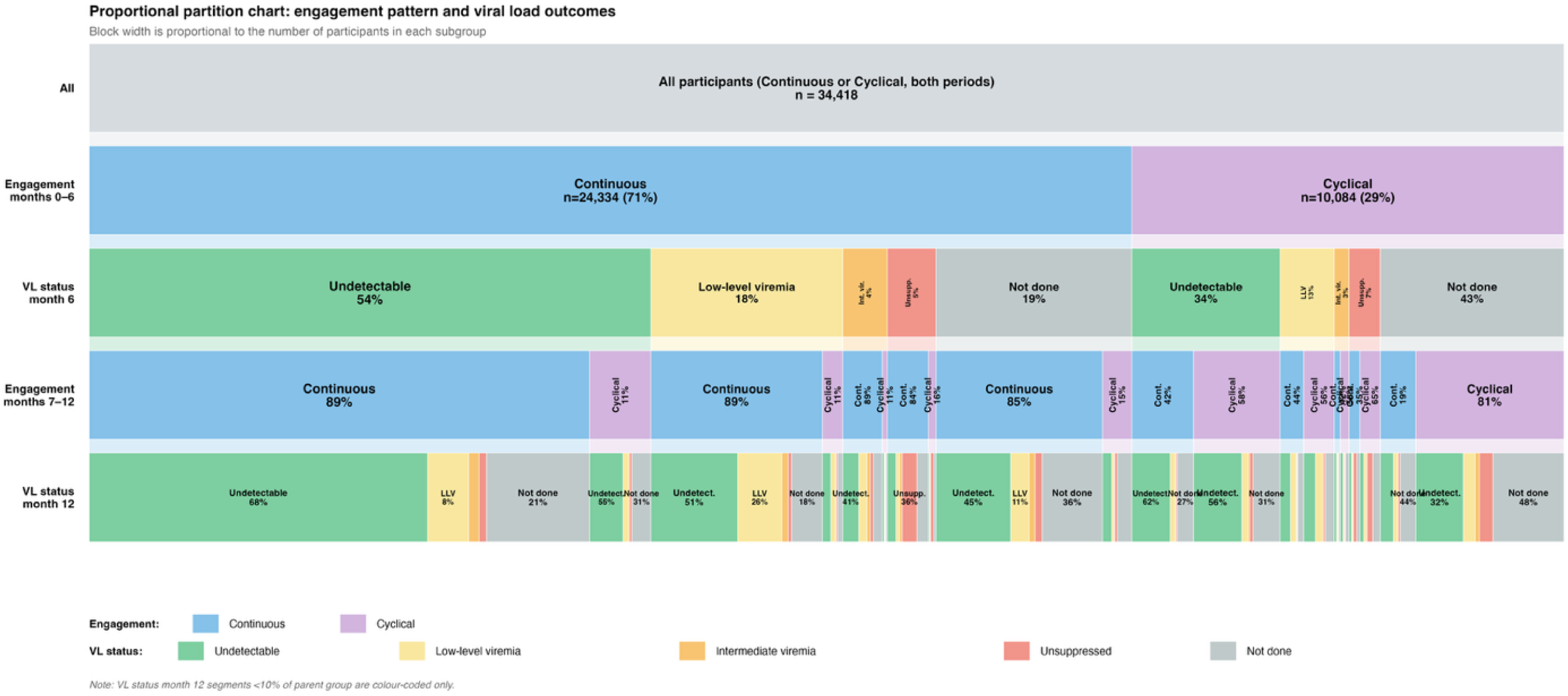
Changes in viral load suppression in the first year among clients retained in care at 12 months

Among the 29% of participants who were cyclically engaged (n=10,084), viral suppression at 6 months was lower (34%), with 13% experiencing LLV, 3% intermediate viremia, 7% unsuppressed, and 43% having no viral load result. By 12 months, 40% were suppressed, 10% had LLV, 3% had intermediate viremia, 8% remained unsuppressed, and 38% had no recorded result. As in the continuously engaged group, the majority of those suppressed at 6 months remained suppressed at 12 months (59%), while 39% of those with LLV at 6 months achieved suppression by 12 months.

### Viral load suppression at 6 and 12 months

In Figure 2 and Supplementary Table 2 we compare 6- and 12-month viral load completion and suppression rates between the two patterns of engagement at each of the respective time points. Within our study sample of clients who remained in care for the 6- or 12-month analytic periods, viral load outcomes improved markedly between 2018 and 2024. Among those with continuous engagement during months 0-6, 6-month viral suppression (<50 copies/mL) increased from 37% in 2018 to 66% in 2024, while the proportion with LLV (50–199 copies/mL) declined from 30% to 7%. Among those with cyclical engagement, 6-month VL suppression increased from 22% to 44% over the same period, though rates remained substantially lower than in the continuous group. Similar patterns were observed at 12 months: among those continuously engaged during months 7-12, suppression increased from 45% in 2018 to 62% in 2024, compared with an increase from 33% to 46% in the cyclical group. Unsuppressed viral loads (≥1000 copies/mL) declined from 5% in 2018 to 2% in 2024. VL testing gaps (“not done”) remained high, particularly in the cyclical group (42% at 6 months vs. 22% for the continuous group; 41% at 12 months vs. 27% for continuous).

**Figure 2.**
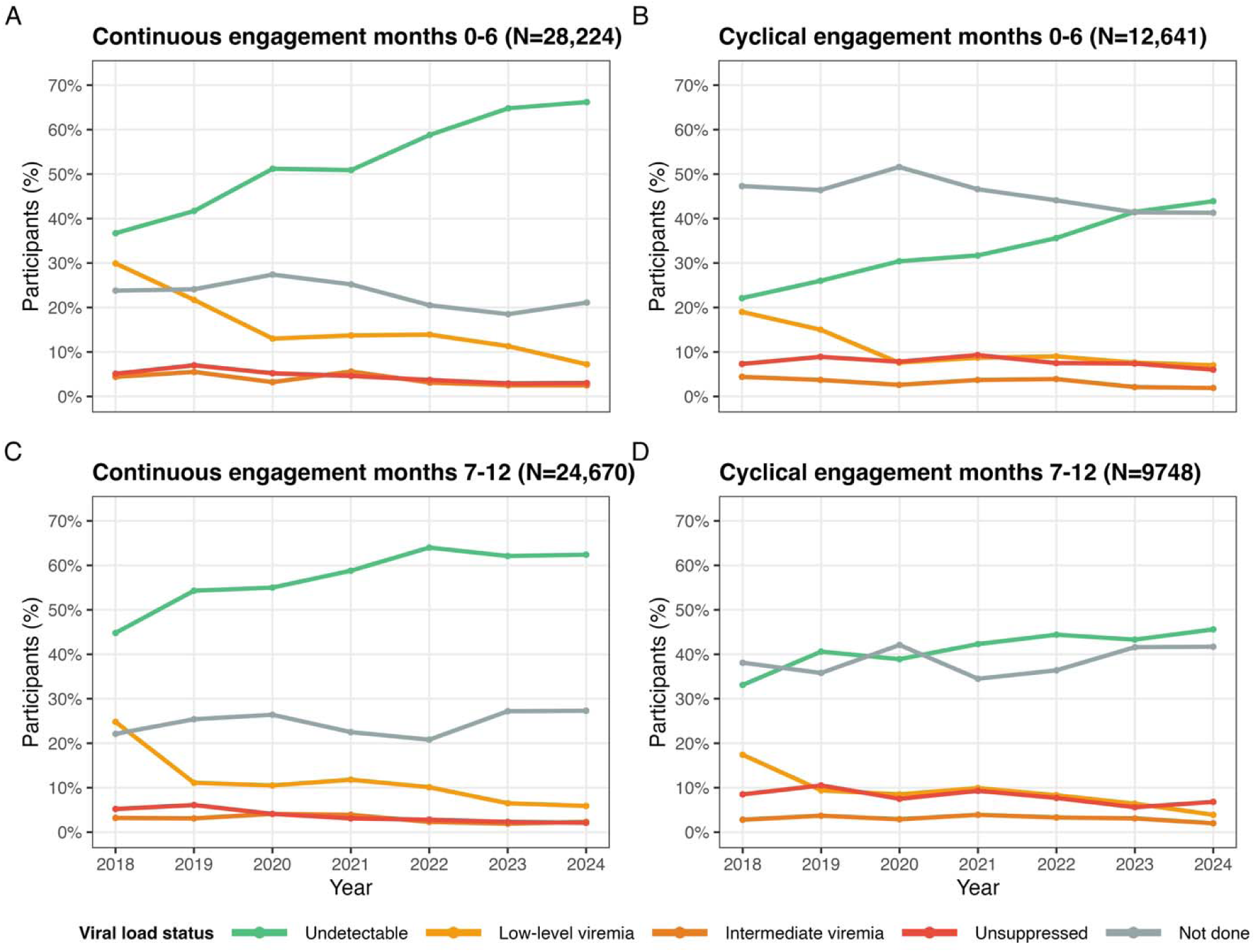
Viral load outcomes by year, time point and engagement pattern

When limiting the denomitator to those who only had a documented viral load test at 6 months (Supplementary Table 3), we observe that the proportion of clients who were virally suppressed remained consistently higher among those engaged in a continuous pattern (48% in 2018 to 84% in 2024) when compared to those enaged in a cyclical pattern (42% in 2018 to 75% in 2024).

### Timing of viral load tests

The timing and type of visit at which viral load tests were conducted, by viral load suppression category and pattern of engagement, are presented in Supplementary Table 3. Among those with suppressed viral loads, the visit at which the viral load was conducted was most likely to have been attended as planned (on time). Clients who were unsuppressed and cyclically engaged were most likely to have had their viral load test performed at a visit following a treatment interruption (i.e. after they were >28 days late).

Figure 3 illustrates the timing of viral load test by visit type. While the timing of viral load testing had an approximately symmetric distribution among those who were continuous, those who were unsuppressed and in a cyclical pattern showed a skewed to the left curve, suggesting that treatment interruption also led to delays in viral load testing and that being unsuppressed was not due to being tested too early. Median time to viral load testing at 6 months was 176 days among continuous engagers and 181 days cyclical engagers, while the 12-month viral load testing medians were 364 days and 370 days, respectively.

**Figure 3.**
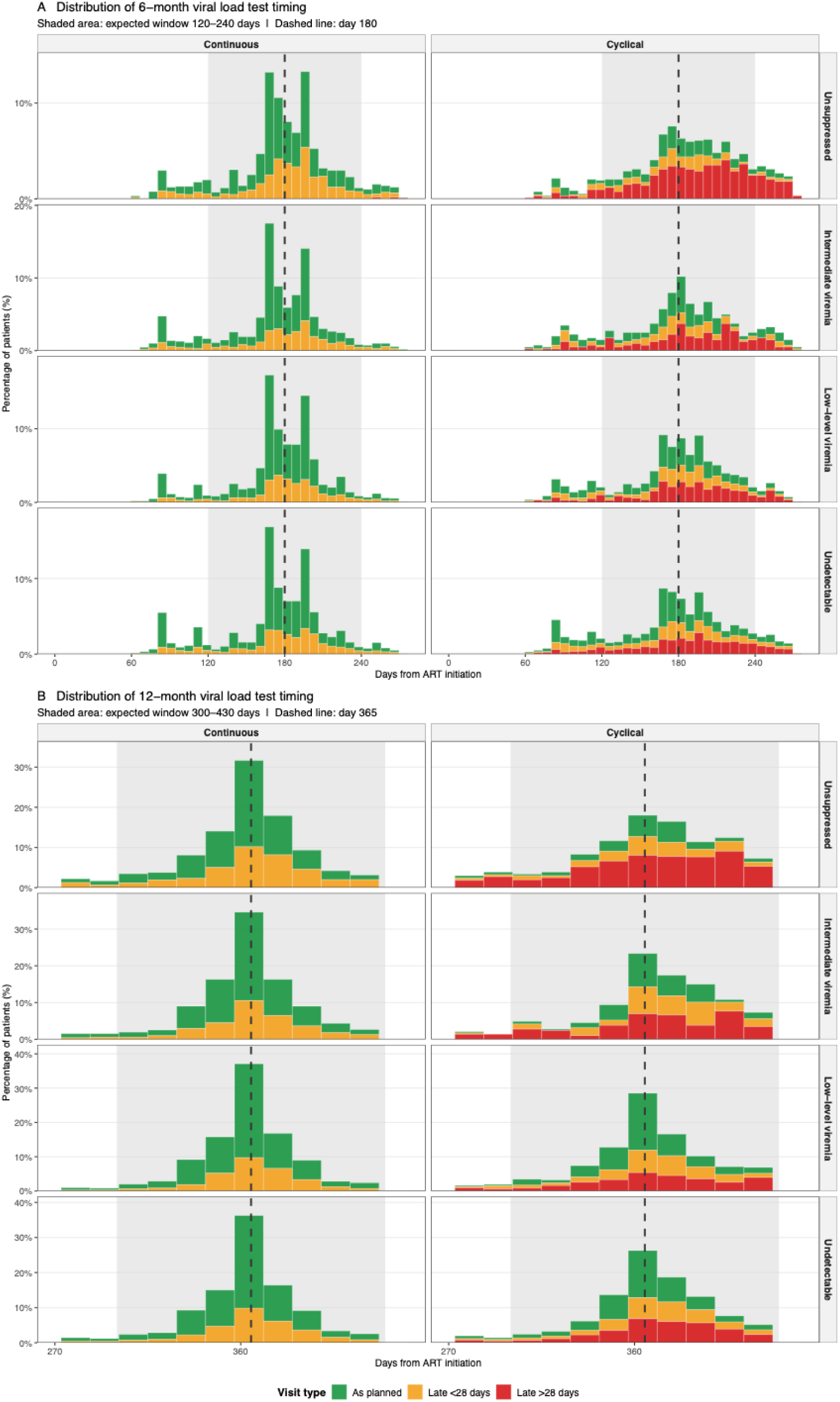
Visit category and timing of viral load testing (percentage of cohort with documented viral load test results)

### Predictors of outcomes

In multivariable modified Poisson regression models, continuous engagement (compared with cyclical engagement) was strongly associated with a higher likelihood of suppression at both 6 months (aRR 1.60, 95% CI 1.55-1.64) and 12 months (aRR 1.38, 95% CI 1.34-1.42) (Figure 4a and 4b). Baseline CD4 count ≥200 cells/mm^3^ (vs. <200) was associated with approximately 20% higher suppression at both time points (6 months: aRR 1.21 95% CI1.18-1.25; 12 months: aRR 1.19 95% CI 1.16-1.22).

**Figure 4.**
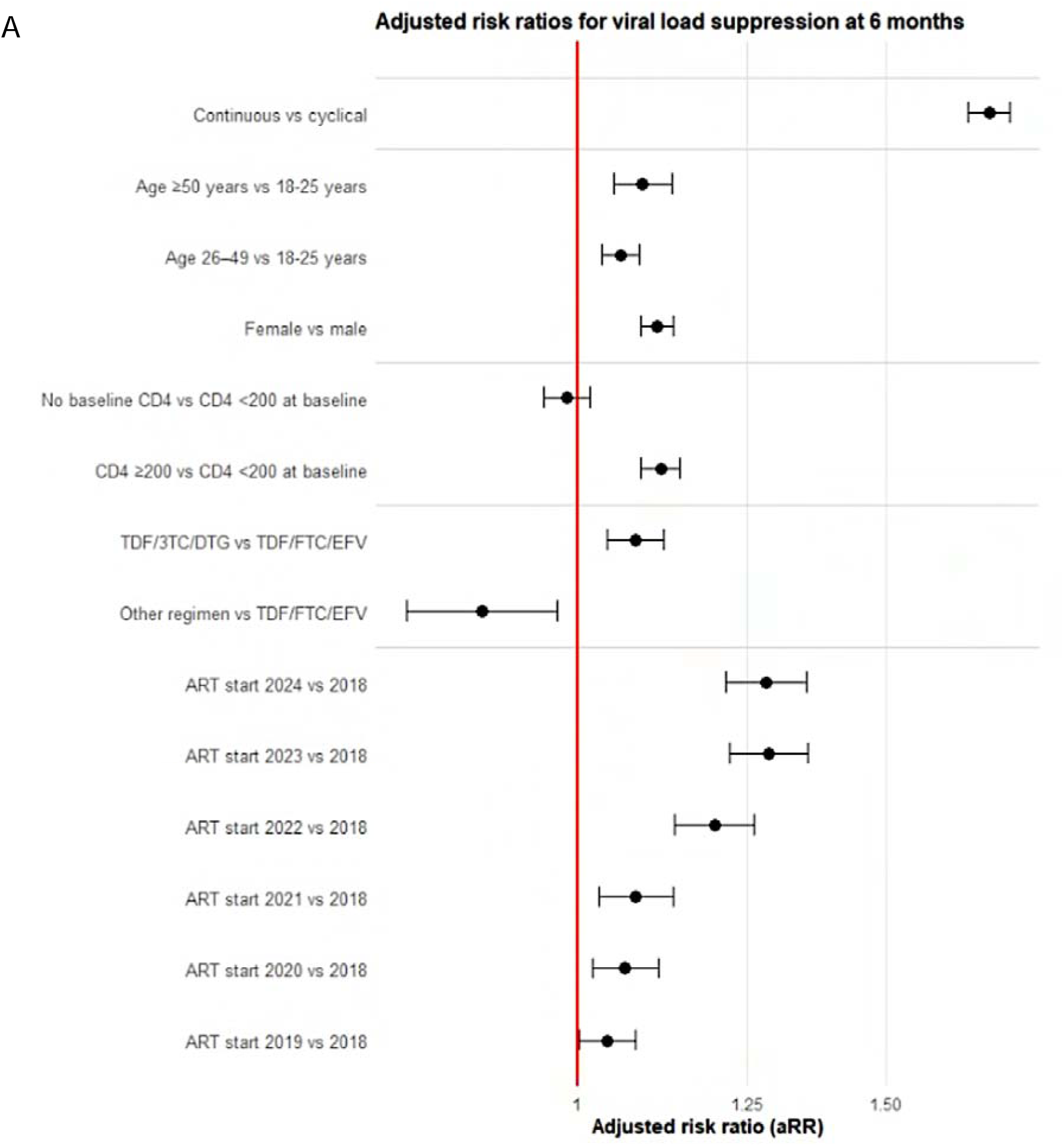
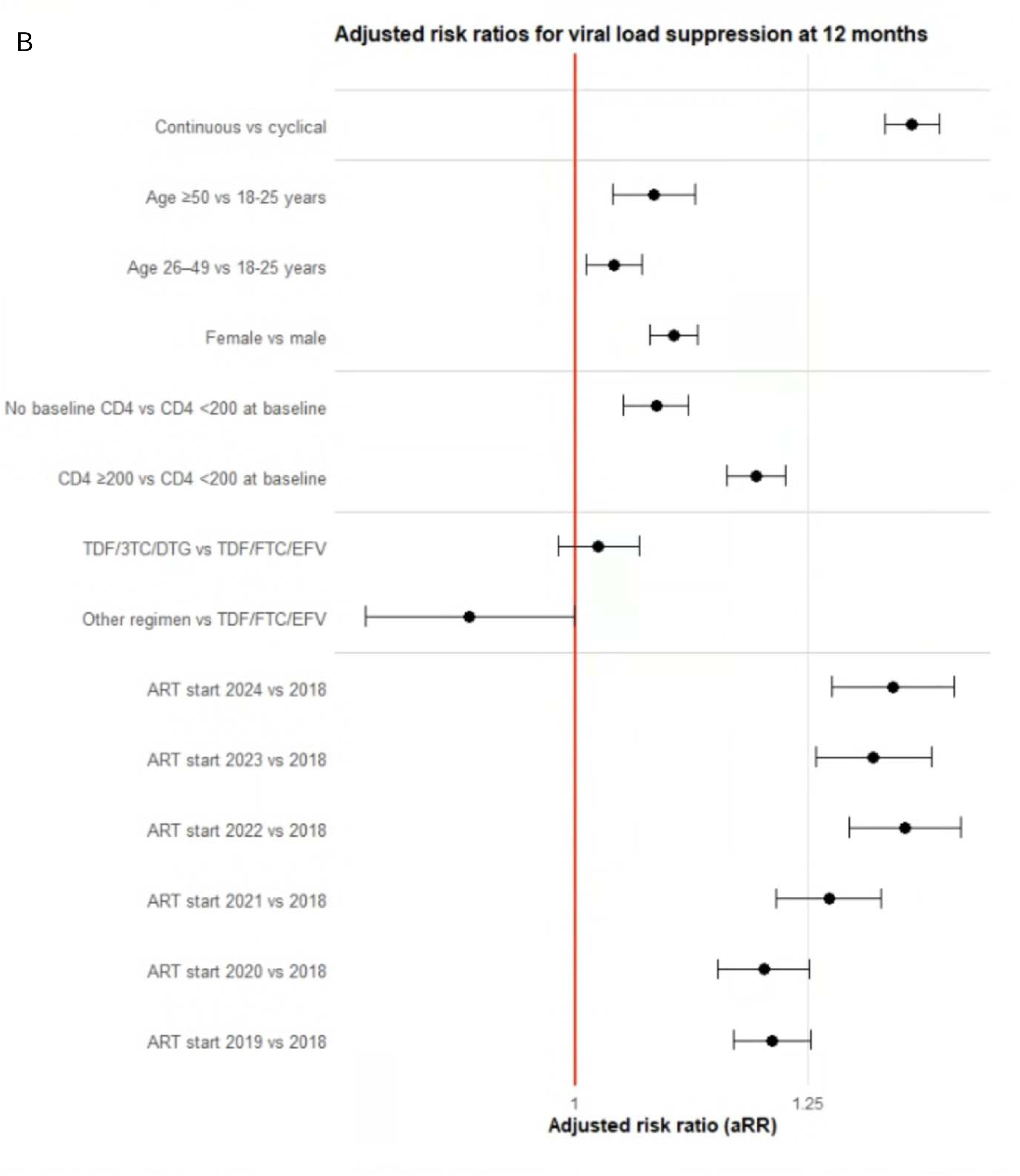
Predictors of viral load suppression A) time of 6-month viral load (N=40,865) and B) time of 12-month viral load (N=34,418) Note: Adjusted risk ratios are derived from a single multivariable model using all variables listed. No other covariates were included in the model.

Calendar year of ART initiation also showed strong association with viral load suppression at both time points. Compared with initiation in 2018, later initiation years were associated with progressively higher suppression. At 6 months, the adjusted risk ratio (aRR) for suppression increased from 1.13 [1.09-1.17] in 2019 to 1.67 [1.57-1.77] in 2024. Twelve-month suppression remained substantially higher in later years as well (2024 vs. 2018: 1.36 [1.28-1.44]; 2022 vs. 2018: 1.53 [1.47-1.60].

Female sex was consistently associated with at least 15% higher suppression than male sex at both time points (6 months: aRR 1.17 [95% CI 1.14-1.20]; 12 months: 1.10 [1.07-1.12]. Those older than 50 had approximately 10% higher likelihood of suppression than those 18-25 years (≥50 years at 6 months: 1.09 [1.05-1.14]; at 12 months: 1.08 [1.04-1.12]. Initiation on a dolutegravir-based regimen (TDF/3TC/DTG vs. TDF/FTC/EFV) was associated with modestly higher 6-month suppression (1.09 [1.04-1.13], but this association attenuated by 12 months (1.02 [0.98-1.06].

In the sensitivity analysis limited to those who were initiated on DTG in 2020 or later (n=19,975), the effect size remained substantial (aRR 1.6 (1.54-1.66)) when comparing continuous to cyclical engagers, but the effect of year of initiation was smaller, with an aRR of 1.30 (1.24-1.35) when comparing those who initiated in 2024 to those who initiated in 2020 (Supplementary figure 2).

When replicating the regression using thresholds of <200 copies/mL and <1000 copies/mL (Supplementary Figure 3), we found that the association between pattern of engagement remained robust with an aRR of 1.50 (95% CI 1.46-1.54) for the threshold of 200 copies/mL and 1.46 (95% CI 1.43-1.51) at a threshold of 1000 copies/mL.

## DISCUSSION

In this large cohort of individuals in their first year of antiretroviral therapy (ART), we found that patterns of early engagement were strongly associated with both viral load test uptake and viral suppression outcomes. Although the majority of participants remained continuously engaged in care at both 6 and 12 months after ART initiation, a substantial proportion experienced cyclical engagement, characterised by one or more treatment interruptions >28 days and subsequent re-engagement. Individuals with continuous engagement consistently demonstrated higher rates of viral suppression than those with cyclical engagement, with this gap widening over time. Continuous engagement was associated with superior virological outcomes across all calendar years, with suppression rates among those continuously engaged improving from 37% in 2018 to 66% in 2024, compared to 22% suppressed in 2018 and 44% in 2024 among cyclical engagers. Among those retained at 12 months over the full six-year dataset, just over half of continuous engagers were virally suppressed, compared with fewer than 40% of cyclical engagers. (We note these values are lower than nationally reported suppression rates because our denominator includes those who did not have a documented viral load test, while most national estimates report suppression among those who had a test.)

Our results demonstrate that cyclical engagement is not merely a marker of imperfect adherence but is associated with clinically meaningful deficits in viral suppression. This finding has important implications at both the individual and programmatic levels, as poorer viral suppression increases the risk of HIV-related morbidity and onward transmission while also undermining progress toward population-level treatment targets. The duration and frequency of short treatment interruptions, even for those who do return for care after interruptions, thus merit more serious attention than they have so far received.

Early cyclical engagers may represent an important sub-population for intensified clinical and adherence support. Not only are they at highest risk of treatment interruptions, which is associated with increased mortality after return to care(5,16,17). These effects may even persist after viral suppression is achieved(18). Importantly, not having a suppressed viral load in the early treatment period is associated with higher risk of non-suppression at 12 months(19).

Our findings are consistent with those reported by Moolla et al, who, using data from South Africa’s Western Cape Province only, reported that treatment interruptions of a median of 103 days were associated with a 3.5- to 4.5-fold increased odds of viral non-suppression over a median of 5 years after ART initiation(6). A national analysis, using laboratory data only and a minimum of 12 months on ART found that missing a prior scheduled viral load test—which might indicate an interruption in care—was highly predictive of non-suppression(20). Neither of these studies, however, reported the association of treatment interruptions and viral suppression during the early treatment period.

For those who had a viral load test in our study, the median days from initiation to testing was nearly identical across all suppression categories and engagement patterns (range 176-184 days). The visit type at which those tests occurred, however, differed by pattern of engagement: continuous engagers had two thirds of viral load tests done at visits attended as planned, while cyclical engagers, whether or not suppressed, most frequently had viral load tests done at visits after a treatment interruption. More than half of unsuppressed individuals with cyclical engagement patterns had their VL tests conducted following prolonged (>28 days) treatment interruptions, highlighting the clinical consequences of disrupted care. Importantly, among all those who were unsuppressed, regardless of engagement pattern, only 47% completed their VL at a visit that was attended as planned, signalling that even interruptions shorter than 28 days may be detrimental to outcomes. More encouragingly, VL test non-completion declined steadily over successive years, reflecting improvements in programme performance, with the notable exception of 2020, when peak levels of missed VL testing occurred, likely due to delayed clinic visit scheduling related to the COVID-19 pandemic. Nevertheless, incomplete VL monitoring remained common, with approximately half of all participants, including 21% of those in care continuously, lacking documented VL results at 6 months and 27% at 12 months in 2024. While there were no differences in age and sex between those with and without a documented VL test, another recent study conducted at the same South African facilities suggests that those who were less educated, not living with a partner, or were not able to obtain cash for healthcare were more likely not to have a documented VL (21).

As anticipated (22), use of dolutegravir-containing regimens was associated with improved suppression at 6 months. This effect diminished by 12 months, however, suggesting that pharmacological potency alone is insufficient to overcome the effects of poor engagement. Continued investment in adherence support and retention strategies is therefore necessary, even in the context of highly effective regimens. Other risk factors for poor virological outcomes included male sex, younger age, and lower baseline CD4 counts, all well established predictors of non-suppression(23,24).

On a more positive note, along with lower rates of VL test non-completion, clients initiating ART in more recent years, particularly in 2023, had substantially higher viral suppression rates. These likely resulted from an improved drug regimen, as noted above, but may also reflect ongoing improvements in programme quality and treatment delivery following revisions to treatment guidelines in 2023 that allowed enrollment in differentiated models of care (DMOCs) as early as six months after initiation.(25) This is supported by our sensitivity analysis, where we found that even when restricting analysis to clients who initiated on a DTG-containing regimen, the effect of year of initiation persisted, albeit to a lesser extent. Since South Africa’s DMOCs allow clients to access medication closer to home and require fewer clinic visits, early enrollment eligibility may also have encouraged better suppression.

The results of this analysis should be interpreted within the context of the limitations of this study and, more broadly, relevant to observational cohorts from routinely collected EMR sources. First, as noted, the datasets analyzed are derived from TIER.Net, which is not networked among facilities. This means that unofficial (“silent”) transfers of ART clients from one facility to another are reflected in the database as loss to follow up (non-retention)(26). Viral load tests conducted at a facility other than the initiating site are not captured in our dataset, and our analysis thus likely underestimates retention and observed viral load testing. Prior analysis quantifying misclassification bias in these datasets due to silent transfers suggests that outcome misclassification occurring in this way would underestimate the risk of disengagement by less than 20 percentage points, however, regardless of the extent of misclassification (5). Recent data indicate that in 2022, 99.1% of viral loads performed by the National Health Laboratory Service were captured in TIER.Net, with the highest capture rates observerd for viral loads <50copies/mL(27). This high level of data completeness, particularly for suppressed viral loads, substantially minimizes the risk of misclassifying virally suppressed individuals as unsuppressed due to missing results.

Second, visit attendance is used here as a proxy for medication possession and treatment adherence. To the extent to which visit attendance occurred without adherence to medication, our estimates over-estimate engagement in care. Conversely, clients who have been on treatment for some time may still have extra medications in hand after missing a clinic visit, and borrowing from other ART patients is also possible(8,21). In this case our estimates underestimate continuous engagement. We note that having extra medications on hand is less likely during the early treatment period as clients have not had sufficient time to build up a medication reserve. Given that the proportion of clients who present for care initiation with prior treatment experience is estimated to greater than 50%, however, it is still possible(8,9).

Third, missingness of viral load testing results is interpreted here as viral load test not being done. It is possible that tests were done but not captured in the EMR, however, or that data text formatting of suppressed test results caused these results to be excluded during data exports. To the extent which this occurred, our analysis underestimates rates of VL test monitoring and, where relevant, viral load suppression. It is likely that this misclassification would be non-differential, however, and thus the impact on our estimates of the effect of pattern of engagement on viral load suppression would be negligible.

Finally, while the dataset here is large (n >40,000) and included diverse facilities in terms of size and setting, we acknowledge that it is limited to 24 clinics in four provinces and may not be generalizable to all settings in South Africa or the region.

## CONCLUSION

Concern about disengagement from HIV care has traditionally focused on long or permanent disruptions, leading either to “re-initiation” of ART or, ultimately, to mortality. We found that even the shorter interruptions that are very frequently observed among ART clients in South Africa are associated with serious effects on health outcomes. As noted above, the duration and frequency of short treatment interruptions thus merit more serious attention than they have so far received. Patterns of engagement in the early treatment period in particular may provide a valuable, routinely measurable indicator of long-term treatment success.

Incorporating engagement metrics into routine monitoring and evaluation frameworks could enable earlier identification of patients at risk of poor outcomes and facilitate targeted, proactive interventions. In the South African public sector, leveraging such data-driven approaches may contribute to more efficient use of limited resources and improved population-level virological control. Growing numbers of clients experiencing cyclical patterns of engagement in HIV care will likely require service delivery approaches adapted to address the drivers underlying treatment interruption, support re-engagement in care and promote re-establishment of adherence among those returning after an interruption. These may include both early provision of multi-month dispensing and early eligibility for differentiated service delivery models that promote continuity of care, including community- and workplace-based ART delivery, adherence clubs, flexible clinic hours, and male and youth-friendly services.

## Authors’ contributions

MB conceptualized the study, analyzed the data, and contributed to writing the original draft of the manuscript. MM1 conceptualized the study and contributed to writing the original draft of the manuscript. BN, NS and AJM contributed to the analysis. AH curated the data and contributed to the analysis. LM and MM2 contextualized the results. SR conceptualized the study, contributed to the analysis, and contributed to writing the original draft of the manuscript. All authors reviewed and edited the final manuscript.

## Data availability

Data used in this study are owned by the South Africa National Department of Health and cannot be shared by the authors. Data can be requested through the National Health Research Database at https://nhrd.health.gov.za/ and contacting each province’s representative as shown.

