## Supplementary figures and images for "The association between HIV treatment interruptions and viral suppression during the early treatment period: retrospective cohort study in South Africa"

### Supplementary figure 1

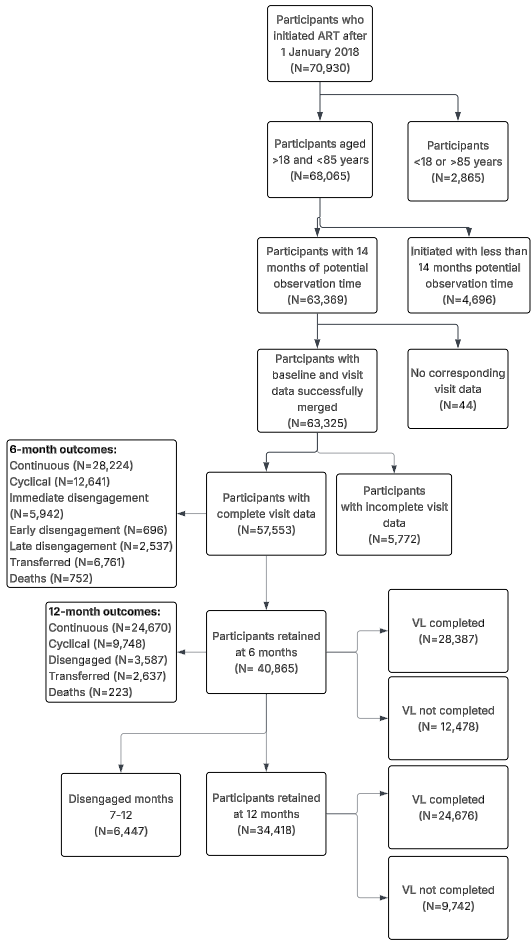

### Supplementary figure 2

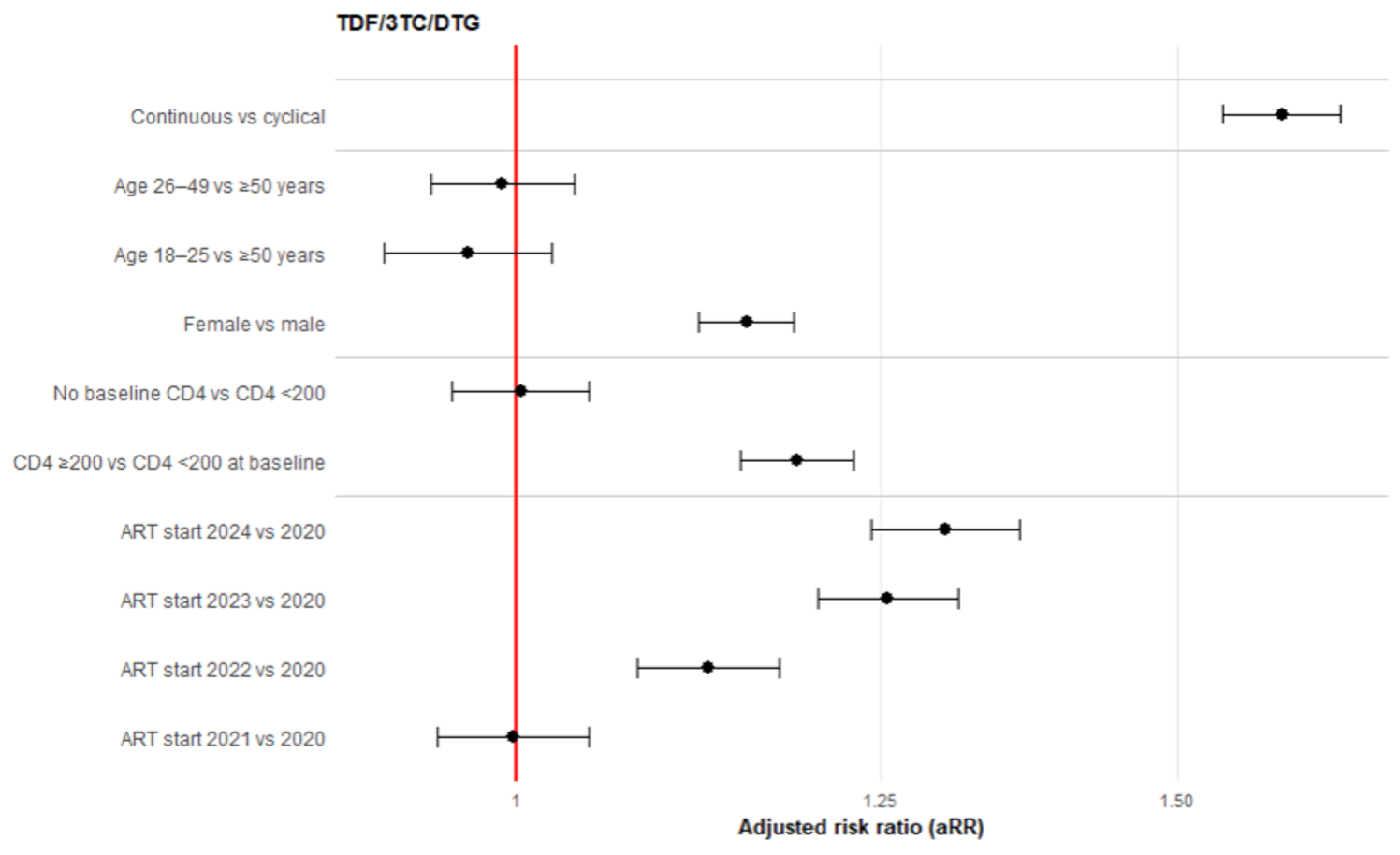

### Supplementary figure 3

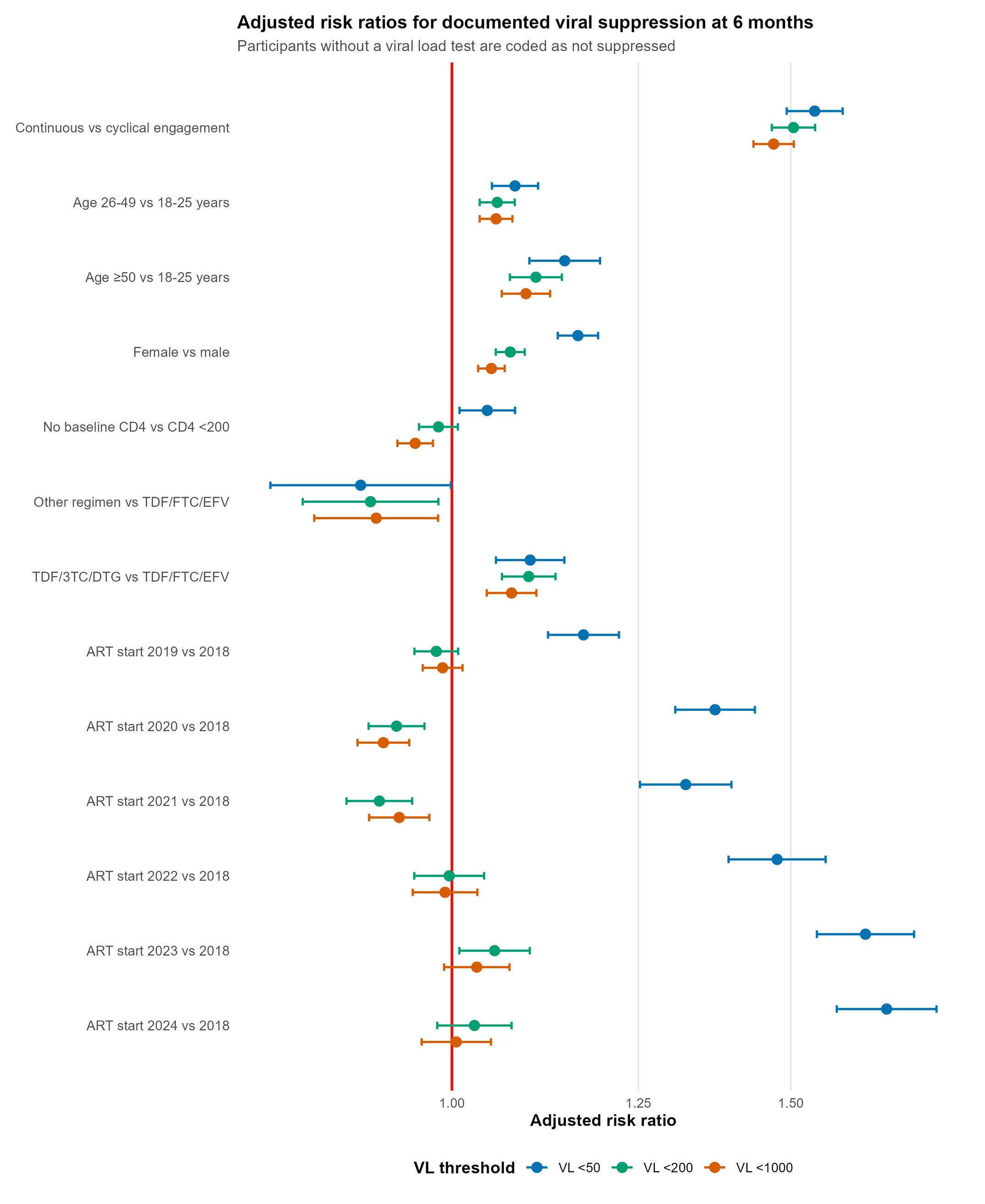
