## Supplementary tables for "The association between HIV treatment interruptions and viral suppression during the early treatment period: retrospective cohort study in South Africa"

Supplementary table 1. Results of total sample

| | Outcome Months 0-6 | | | | | | | |
|---|---|---|---|---|---|---|---|---|
| | Continuous (N=28224) | Cyclical (N=12641) | Death (N=752) | Early (N=696) | Immediate (N=5942) | Late (N=2537) | TFO (N=6761) | Overall (N=57553) |
| Outcome Months 7-12 | | | | | | | | |
| Continuous | 21447 (76.0%) | 3223 (25.5%) | 0 (0%) | 0 (0%) | 0 (0%) | 0 (0%) | 0 (0%) | 24670 (42.9%) |
| Cyclical | 2887 (10.2%) | 6861 (54.3%) | 0 (0%) | 0 (0%) | 0 (0%) | 0 (0%) | 0 (0%) | 9748 (16.9%) |
| Disengaged months 0-6 | 0 (0%) | 0 (0%) | 0 (0%) | 696 (100%) | 5942 (100%) | 2537 (100%) | 0 (0%) | 9175 (15.9%) |
| Disengaged months 7-12 | 2115 (7.5%) | 1472 (11.6%) | 0 (0%) | 0 (0%) | 0 (0%) | 0 (0%) | 0 (0%) | 3587 (6.2%) |
| Transferred | 1638 (5.8%) | 999 (7.9%) | 0 (0%) | 0 (0%) | 0 (0%) | 0 (0%) | 6761 (100%) | 9398 (16.3%) |
| Death | 137 (0.5%) | 86 (0.7%) | 752 (100%) | 0 (0%) | 0 (0%) | 0 (0%) | 0 (0%) | 975 (1.7%) |

**Supplementary Table 2. Viral load suppression by pattern of engagement and year of initiation**

| Suppression category | 2018 | 2019 | 2020 | 2021 | 2022 | 2023 | 2024 |
|---|---|---|---|---|---|---|---|
| | **6-month viral load** | | | | | | |
| | *Continuous engagement months 0-6 (n=28,224)* | | | | | | |
| N | (N=4714) | (N=6336) | (N=4905) | (N=4008) | (N=3384) | (N=2869) | (N=2008) |
| Not done | 1123 (24%) | 1526 (24%) | 1346 (27%) | 1011 (25%) | 694 (21%) | 532 (19%) | 424 (21%) |
| Unsuppressed | 241 (5%) | 442 (7%) | 253 (5%) | 184 (5%) | 124 (4%) | 83 (3%) | 60 (3%) |
| Intermediate viremia | 208 (4%) | 349 (6%) | 155 (3%) | 223 (6%) | 106 (3%) | 72 (3%) | 50 (2%) |
| Low-level viremia | 1410 (30%) | 1376 (22%) | 639 (13%) | 549 (14%) | 469 (14%) | 324 (11%) | 145 (7%) |
| Undetectable | 1732 (37%) | 2643 (42%) | 2512 (51%) | 2041 (51%) | 1991 (59%) | 1858 (65%) | 1329 (66%) |
| | *Cyclical engagement months 0-6 (n=12,641)* | | | | | | |
| N | (N=2684) | (N=2502) | (N=1778) | (N=1621) | (N=1578) | (N=1455) | (N=1023) |
| Not done | 1269 (47%) | 1160 (46%) | 917 (52%) | 756 (47%) | 696 (44%) | 602 (41%) | 422 (41%) |
| Unsuppressed | 195 (7%) | 222 (9%) | 138 (8%) | 150 (9%) | 118 (7%) | 107 (7%) | 61 (6%) |
| Intermediate viremia | 117 (4%) | 93 (4%) | 47 (3%) | 60 (4%) | 61 (4%) | 31 (2%) | 19 (2%) |
| Low-level viremia | 510 (19%) | 376 (15%) | 135 (8%) | 141 (9%) | 142 (9%) | 111 (8%) | 72 (7%) |
| Undetectable | 593 (22%) | 651 (26%) | 541 (30%) | 514 (32%) | 561 (36%) | 604 (42%) | 449 (44%) |
| | **12-month viral load** | | | | | | |
| | *Continuous engagement months 7-12 (n=24,670)* | | | | | | |
| N | (N=4247) | (N=5467) | (N=4236) | (N=3446) | (N=2956) | (N=2582) | (N=1736) |
| Not done | 937 (22%) | 1391 (25%) | 1117 (26%) | 776 (23%) | 616 (21%) | 703 (27%) | 474 (27%) |
| Unsuppressed | 219 (5%) | 331 (6%) | 173 (4%) | 106 (3%) | 83 (3%) | 59 (2%) | 37 (2%) |
| Intermediate viremia | 134 (3%) | 168 (3%) | 172 (4%) | 133 (4%) | 67 (2%) | 48 (2%) | 40 (2%) |
| Low-level viremia | 1054 (25%) | 609 (11%) | 445 (11%) | 406 (12%) | 299 (10%) | 168 (7%) | 102 (6%) |
| Undetectable | 1903 (45%) | 2968 (54%) | 2329 (55%) | 2025 (59%) | 1891 (64%) | 1604 (62%) | 1083 (62%) |
| | *Cyclical engagement months 7-12 (n=9,748)* | | | | | | |
| N | (N=1964) | (N=1875) | (N=1460) | (N=1323) | (N=1257) | (N=1056) | (N=813) |
| Not done | 749 (38%) | 672 (36%) | 615 (42%) | 457 (35%) | 457 (36%) | 439 (42%) | 339 (42%) |
| Unsuppressed | 167 (9%) | 197 (11%) | 110 (8%) | 123 (9%) | 97 (8%) | 59 (6%) | 55 (7%) |
| Intermediate viremia | 55 (3%) | 69 (4%) | 43 (3%) | 52 (4%) | 41 (3%) | 33 (3%) | 16 (2%) |
| Low-level viremia | 342 (17%) | 176 (9%) | 124 (8%) | 131 (10%) | 104 (8%) | 68 (6%) | 32 (4%) |
| Undetectable | 651 (33%) | 761 (41%) | 568 (39%) | 560 (42%) | 558 (44%) | 457 (43%) | 371 (46%) |

### Supplementary table 3. Viral load suppression among those with viral loads by pattern of engagement and year of initiation

| Suppression category | 2018 | 2019 | 2020 | 2021 | 2022 | 2023 | 2024 |
|---|---|---|---|---|---|---|---|
| | | | | **6-month viral load** | | | |
| | | | | *Continuous engagement months 0-6 (n=21,568)* | | | |
| N | (N=3591) | (N=4810) | (N=3559) | (N=2997) | (N=2690) | (N=2337) | (N=1584) |
| Unsuppressed | 241 (7%) | 442 (9%) | 253 (7%) | 184 (6%) | 124 (5%) | 83 (4%) | 60 (4%) |
| Intermediate viremia | 208 (6%) | 349 (7%) | 155 (4%) | 223 (7%) | 106 (4%) | 72 (3%) | 50 (3%) |
| Low-level viremia | 1410(39%) | 1376(29%) | 639 (18%) | 549 (18%) | 469 (17%) | 324 (14%) | 145 (9%) |
| Undetectable | 1732 (48%) | 2643 (55%) | 2512 (71%) | 2041 (68%) | 1991 (74%) | 1858 (80%) | 1329 (84%) |
| | | | | *Cyclical engagement months 0-6 (n=6,819))* | | | |
| N | (N=1415) | (N=1342) | (N=861) | (N=865) | (N=882) | (N=853) | (N=601) |
| Unsuppressed | 195 (14%) | 222 (17%) | 138 (16%) | 150 (17%) | 118 (13%) | 107 (13%) | 61 (10%) |
| Intermediate viremia | 117 (8%) | 93 (7%) | 47 (5%) | 60 (7%) | 61 (7%) | 31 (4%) | 19 (3%) |
| Low-level viremia | 510 (36%) | 376 (28%) | 135 (16%) | 141 (16%) | 142 (16%) | 111 (13%) | 72 (12%) |
| Undetectable | 593 (42%) | 651 (49%) | 541 (63%) | 514 (59%) | 561 (64%) | 604 (71%) | 449 (75%) |
| | | | | **12-month viral load** | | | |
| | | | | *Continuous engagement months 7-12 (n=18,656)* | | | |
| N | (N=3310) | (N=4076) | (N=3119) | (N=2670) | (N=2340) | (N=1879) | (N=1262) |
| Unsuppressed | 219 (7%) | 331 (8%) | 173 (6%) | 106 (4%) | 83 (4%) | 59 (3%) | 37 (3%) |
| Intermediate viremia | 134 (4%) | 168 (4%) | 172 (6%) | 133 (5%) | 67 (3%) | 48 (3%) | 40 (3%) |
| Low-level viremia | 1054(32%) | 609 (15%) | 445 (14%) | 406 (15%) | 299 (13%) | 168 (9%) | 102 (8%) |
| Undetectable | 1903(57%) | 2968(73%) | 2329(75%) | 2025(76%) | 1891(81%) | 1604(85%) | 1083(86%) |
| | | | | *Cyclical engagement months 7-12 (n=6,020)* | | | |
| Not done | (N=1215) | (N=1203) | (N=845) | (N=866) | (N=800) | (N=617) | (N=474) |
| Unsuppressed | 167 (14%) | 197 (16%) | 110 (13%) | 123 (14%) | 97 (12%) | 59 (10%) | 55 (12%) |
| Intermediate viremia | 55 (5%) | 69 (6%) | 43 (5%) | 52 (6%) | 41 (5%) | 33 (5%) | 16 (3%) |
| Low-level viremia | 342 (28%) | 176 (15%) | 124 (15%) | 131 (15%) | 104 (13%) | 68 (11%) | 32 (7%) |
| Undetectable | 651 (54%) | 761 (63%) | 568 (67%) | 560 (65%) | 558 (70%) | 457 (74%) | 371 (78%) |

**Supplementary table 4. Visit category and timing of viral load testing***

| Status at 6 month viral load test | | | | | | | | |
|---|---|---|---|---|---|---|---|---|
| | **Suppressed** | | **Low level viremia** | | **Intermediate Viremia** | | **Unsuppressed** | |
| **Variable** | **Continuous** | **Cyclical** | **Continuous** | **Cyclical** | **Continuous** | **Cyclical** | **Continuous** | **Cyclical** |
| N | 13422 (79%) | 3588 (21%) | 4671 (77%) | 1369 (23%) | 1096 (73%) | 402 (27%) | 1312 (58%) | 941 (42%) |
| *Visit type* | | | | | | | | |
| As planned | 9286 (69%) | 1533 (43%) | 3307 (71%) | 569 (42%) | 749 (68%) | 156 (39%) | 809 (61.4%) | 245 (26%) |
| Late <28 days | 4135 (31%) | 895 (25%) | 1364 (29%) | 331 (24%) | 347(32%) | 90 (22%) | 503 (38.2%) | 157 (17%) |
| Late >28 days | 0 (0.0%) | 1150 (32%) | 0 (0.0%) | 469 (34%) | 0 (0.0%) | 156 (39%) | 0 (0.0%) | 539 (57%) |
| *Timing of 6-month viral load (days)* | | | | | | | | |
| Median [Q1, Q3] | 176 [161, 196] | 182 [162, 206] | 178 [166, 196] | 182 [159,206] | 176 [162,196] | 182 [154,211] | 181 [165, 198] | 189 [163, 219] |
| **Status at 12 month viral load test** | | | | | | | | |
| | **Suppressed** | | **Low level viremia** | | **Intermediate Viremia** | | **Unsuppressed** | |
| **Variable** | **Continuous** | **Cyclical** | **Continuous** | **Cyclical** | **Continuous** | **Cyclical** | **Continuous** | **Cyclical** |
| N | 12,773 (78%) | 3,690 (22%) | 2901 (76%) | 933 (24%) | 711 (71%) | 287 (29%) | 954 (55%) | 772 (45%) |
| *Visit type* | | | | | | | | |
| As planned | 8,741 (69%) | 1,510 (41%) | 2006 (69%) | 404 (43%) | 470 (66%) | 83 (29%) | 576 (60%) | 161 (21%) |
| Late <28 days | 4,031(32%) | 940 (26%) | 895 (31%) | 248 (27%) | 241 (34%0 | 83 (29%) | 378 (40%) | 156 (20%) |
| Late >28 days | 0 (0.0%) | 1,240 (34%) | 0 (0.0%) | 281 (30%) | 0 (0.0%) | 121 (42%) | 0 (0.0%) | 455 (59%) |
| *Timing of 12-month viral load (days)* | | | | | | | | |
| Median [Q1, Q3] | 364 [352, 376] | 369 [355, 386] | 364 [353, 376] | 366 [354,385] | 364 [351,377] | 372 [356,391] | 365 [350, 378] | 370 [347, 392] |

*Among those who were engaged at each time point and had a viral load N=28,387 for 6-month viral load and N=24,676 for 12-month viral load.
